# Saliva Sample as a Non-Invasive Specimen for the Diagnosis of Coronavirus Disease-2019 (COVID-19): a Cross-Sectional Study

**DOI:** 10.1101/2020.04.17.20070045

**Authors:** Ekawat Pasomsub, Siriorn P. Watcharananan, Kochawan Boonyawat, Pareena Janchompoo, Garanyuta Wongtabtim, Worramin Suksuwan, Somnuek Sungkanuparph, Angsana Phuphuakrat

**Affiliations:** Department of Pathology, Faculty of Medicine Ramathibodi Hospital, Mahidol University, Bangkok, Thailand; Department of Medicine, Faculty of Medicine Ramathibodi Hospital, Mahidol University, Bangkok, Thailand; Chakri Naruebodindra Medical Institute, Faculty of Medicine Ramathibodi Hospital, Mahidol University, Samut Prakan, Thailand

**Author notes:** **Corresponding author:** Angsana Phuphuakrat, MD, PhD, Assistant Professor, Department of Medicine, Faculty of Medicine Ramathibodi Hospital, Mahidol University, 270 Rama VI Rd., Bangkok 10400, Thailand.

**Keywords:** COVID-19, nasopharyngeal swab, RT-PCR, saliva, SARS-CoV-2, throat swab

## Abstract

**Objectives:** Amid the increasing number of global pandemic coronavirus disease 2019 (COVID-19) cases, there is a need for a quick and easy method to obtain a non-invasive sample for the detection of this novel coronavirus 2019 (SARS-CoV-2). We aimed to investigate the potential use of saliva samples as a non-invasive tool for the diagnosis of COVID-19.

**Methods:** From 27 March to 4 April, 2020, we prospectively collected saliva samples and a standard nasopharyngeal and throat swab in persons seeking care at an acute respiratory infection clinic in a university hospital during the outbreak of COVID-19. Real-time polymerase chain reaction (RT-PCR) was performed, and the results of the two specimens were compared.

**Results:** Two-hundred pairs of the samples were collected. Sixty-nine (34.5%) patients were male, and the median (interquartile) age was 36 (28-48) years. Using nasopharyngeal and throat swab RT-PCR as the reference standard, the prevalence of COVID-19 diagnosed by nasopharyngeal and throat swab RT-PCR was 9.5%. The sensitivity and specificity of the saliva sample RT-PCR were 84.2% [95% confidence interval (CI) 79.2%-89.3%], and 98.9% (95% CI 97.5-100.3%), respectively. An analysis of the agreement between the two specimens demonstrated 97.5% observed agreement (kappa coefficient 0.851, 95% CI 0.723-0.979; *p* <0.001).

**Conclusions:** Saliva specimens can be used for the diagnosis of COVID-19. The collection method is non-invasive, and non-aerosol generating. Using a saliva sample as a specimen for the detection of SARS-CoV-2 could facilitate the diagnosis of the disease, which is one of the strategies that helps in controlling the epidemic.

## Introduction

Since December 2019, the outbreak of coronavirus disease 2019 (COVID-19), caused by the novel coronavirus 2019 (SARS-CoV-2) has emerged in Hebei Province of China and has spread to other parts of the world [1, 2]. The number of cases has been increasing rapidly, with a case-fatality rate of 2.3% [3].

Detection of SARS-CoV-2 in patient specimens is the first crucial step for the guidance of treatment, effective infection control in the hospital and control of infection in the community. Screening of infection in suspected cases with a nucleic acid amplification test (NAAT), such as real-time polymerase chain reaction (RT-PCR), in respiratory specimens, is recommended by the World Health Organization [4]. However, the collection of nasopharyngeal and/or oropharyngeal swab specimens is a relatively invasive method and the procedure might put healthcare workers at higher risk for disease transmission during patients’ gag reflex, cough, or sneezing.

As SARS-CoV-2 viral load was demonstrated to present near presentation onset [5], using a saliva sample as the specimen for the screening of the disease is appealing. To determine the potential of using a saliva sample for the diagnosis of COVID-19, we conducted a prospective study investigating the correlation of detection of SARS-CoV-2 in a saliva sample, and nasopharyngeal and throat swabs in patients under investigation at an acute respiratory infection clinic at a university hospital in Bangkok, Thailand during the COVID-19 outbreak.

## Methods

### Study population

A prospective study was conducted among 200 patients under investigation who attended an acute respiratory infection clinic at Ramathibodi Hosptial, Bangkok, Thailand, during 27 March and 4 April, 2020. The inclusion criteria were those who presented with a history of fever or acute respiratory symptoms together with 1) travel history from an endemic area of COVID-19 within 14 days, or 2) contact with an individual who was confirmed or suspected having COVID-19. Patients aged less than 18 years old were excluded.

Patient characteristics, symptoms at presentation, and risk factors were collected. As a standard protocol, nasopharyngeal and throat swabs from a patient were collected using Copan FLOQSwabs® and a sterile tube containing Copan’s Universal Transport Medium™ (UTM®) (Copan Diagnostics). Prior to collecting the swabs, patients were asked to provide a saliva sample, void of coughing, in a sputum collection container containing the UTM®.

The study protocol was reviewed and approved by the Ethical Clearance Committee on Human Right Related to Research Involving Human Subjects of the Faculty of Medicine Ramathibodi Hospital, Mahidol University.

### Specimen processing

Nasopharyngeal and throat swab in a tube and saliva sample from the collection container were treated with the lysis buffer (BioMerieux) to inactivate the SARS-CoV-2. Viral RNA was extracted from 200 μl of the samples within 26 minutes using MagDEA® Dx reagents (Precision System Science) fully automated nucleic acid extraction system, according to the manufacturer’s instructions.

### RT-PCR workflow

The detection of SARS-CoV-2 in the specimens was performed by real-time RT-PCR amplification of SARS-CoV-2 *ORF1AB* and *N* gene fragments, using a SARS-CoV-2 Nucleic Acid Diagnostic Kit (Sansure Biotech) which was approved for detection of the SARS-CoV-2 by the National Medical Products Administration (NMPA) and certified by the China Food and Drug Administration (CFDA) [6]. The lower limit of detection of the test was 200 copies/sample. RT-PCR was performed by the CFX96 Real-Time Detection System (Bio-Rad). The result was considered positive if the cycle threshold (Ct) values of both target genes were ≤38, and negative when Ct values of both targets were >38. Retesting was done among the samples with discordancy of the Ct values; i.e. samples with one target gene with a Ct value of ≤38 and another showing a Ct value of >38. Among the retesting, the specimens with repeated discordancy were reported as negative. The turnaround time of the diagnosis was approximately four hours.

### Statistical analysis

Data were analysed for normality and descriptive statistics presented as a number (percent) for categorical variables and mean±standard deviation (SD) or median (interquartile range; IQR) for continuous variables. Chi-square or Fisher’s exact test was used for categorical variables. Sensitivity, specificity, positive predictive value (PPV), and negative predictive value (NPV) and a 95% confidence interval (CI) were calculated to assess diagnostic performance. The kappa coefficient [7] was used to estimate for the agreement between the saliva RT-PCR and nasopharyngeal and throat swab RT-PCR results. All statistical analyses were performed using Stata statistical software version 15.1 (StataCorp, College Station, TX, 2018).

## Results

Two-hundred pairs of samples of nasopharyngeal and throat swab, and saliva samples were collected. Sixty-nine (34.5%) patients were male. The median (IQR) age was 36 (28-48) years. Median (IQR) onset of symptoms was 3 (2-7) days. Characteristics of the patients are presented in Table 1. The prevalence of COVID-19 diagnosed by nasopharyngeal and throat swab RT-PCR, and saliva RT-PCR in this study were 9.5% and 9.0%, respectively.

**Table 1.**
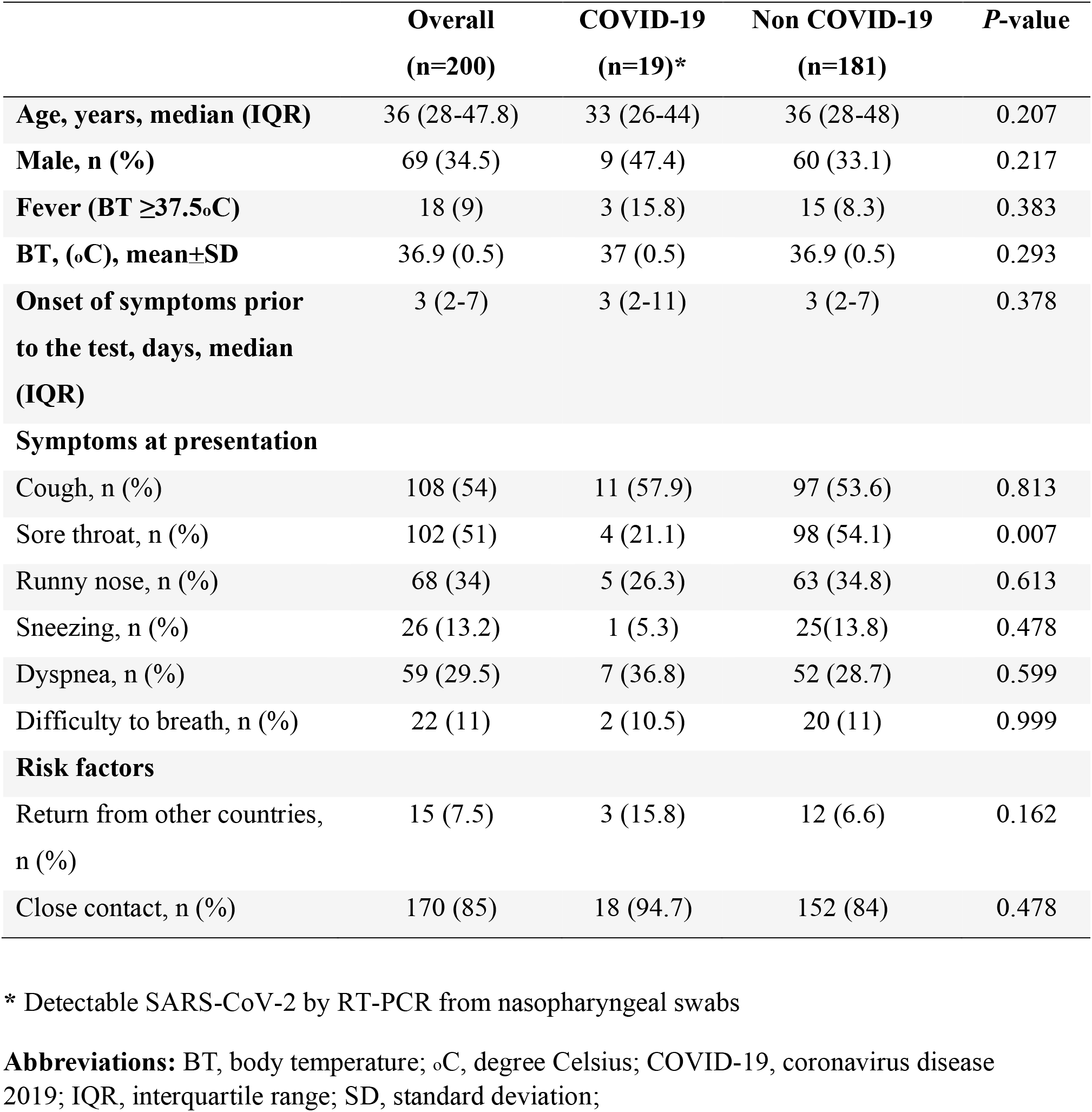
Characteristics of patients under investigation diagnosed with COVID-19 by RT-PCR from nasopharyngeal and throat swabs

Among 19 patients diagnosed with COVID-19 by nasopharyngeal swab RT-PCR, the median age was 33 (26-44) years. Fever as defined by temperature ≥37.5°C was presented in 3 (15.8%) patients. The mean±SD temperature was 37±0.46°C and the mean±SD onset of symptoms prior to the test was 6.35±5.7 days. Common symptoms at presentation were cough (11, 57.9%), dyspnoea (7, 36.8%), and runny nose (5, 26.3%). When compared with 181 patients with negative nasopharyngeal and throat swab RT-PCR, only a sore throat at presentation was significantly lower in the COVID-19 patients. Other characteristics, symptoms at presentation and risk factors were not significantly different (Table 1).

To determine the diagnostic test performance of RT-PCR of the saliva, RT-PCR results of the nasopharyngeal and throat swabs were used as the reference standard. The sensitivity and specificity of saliva samples were 84.2% (95% CI 79.2%-89.3%), and 98.9% (95% CI 97.5-100.3%), respectively (Table 2). PPV and NPV were 88.9% (95% CI 84.5%-93.2%), and 98.4% (95% CI 96.6-100.1%), respectively. An analysis of the agreement between the two specimens revealed a 97.5% observed agreement (kappa coefficient 0.851, 95% CI 0.723-0.979; *p* <0.001).

**Table 2.**
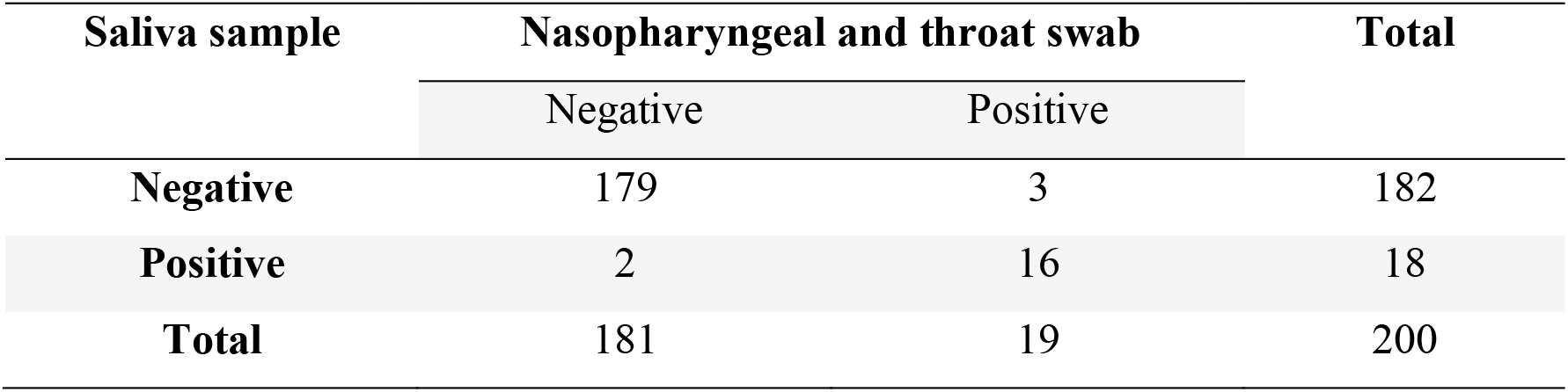
The comparison for the detection of SARS-CoV-2 RT-PCR between nasopharyngeal and throat swab, and saliva sample

## Discussion

The present study showed the value of testing a saliva sample as a non-invasive detection of SARS-CoV-2. The Saliva PCR test demonstrated high sensitivity and comparable performance to the current standard of nasopharyngeal and throat swab. The kappa coefficient value showed a strong agreement of the diagnosis between the standard nasopharyngeal and throat swab and the saliva sample.

Like severe acute respiratory syndrome-coronavirus (SARS-CoV), a causative agent of the severe acute respiratory syndrome (SARS), SARS-CoV-2 employs the host-cell angiotensin-converting enzyme 2 (ACE2) as the main host receptor for cellular entry [8]. Previous experimental studies showed a higher level of ACE2 expression in minor salivary glands, as compared to that in the lungs [9] and that the epithelial cells lining salivary gland duct were early targets cells of SARS-CoV infection in rhesus macaques [10]. SARS-CoV was also detected in saliva samples [11]. This suggested that the salivary glands could be a potential target for SAR-CoV-2 infection, and hence saliva could be a potential sample for SARS-CoV-2 detection.

From recent findings, SARS-CoV-2 was detected from posterior oropharyngeal saliva samples, with a notable high viral load at the disease presentation. [5, 12]. In their protocol, an early morning saliva was collected after coughing up by clearing the throat. In our study, a saliva sample was the patient’s self-generated, without a need for coughing up. This non-invasive procedure might be less aerosol-generating and might reduce the risk of infection for health care workers working in the clinic.

Although testing of saliva might provide an advantage as an easy procedure, a comparison study between saliva and confirmed case of bronchoalveolar lavage (BAL) fluid or convalescence serum titre has not been available. A recent study that detected the virus from multiple sites showed a lower test positivity rate from the nasal swab (63%), as compared to the bronchoalveolar lavage fluid (93%) [13]. Therefore, a false negative test of the SARS-CoV-2 from saliva sample might possible, and this would be an area for further exploration. However, the spectrum of the disease ranges from asymptomatic, upper respiratory tract symptoms, pneumonia, and acute respiratory distress syndrome [14, 15]. Hence, the discrepancy of SARS-CoV-2 detection from different specimens might also be possible.

The study had several strengths. We prospectively collected data on consecutive patients who were at high risk of COVID-19 infection including those with acute respiratory symptoms and having risk factors, thus minimizing potential spectrum effect. All enrolled patients were verified with the reference standard. As for the limitations, two specimens had detectable of SARS-CoV-2 from saliva samples, but were undetectable from nasopharyngeal and throat swab. The significance of these positive results are unknown since we do not have clinical data on any follow-up of these patients.

With the current situation of a shortage supply of personal protective equipment during the pandemic and moderate risk of infection among healthcare workers, a saliva sample is an alternative specimen collection for the diagnosis of COVID-19, especially in resource-limited settings.

## Data Availability

All relevant data are within the paper.

## Conflict of interest

All authors declare that there is no conflicts of interest.

## Funding

This study was supported by a grant from Faculty of Medicine Ramathibodi Hospital, Mahidol University.

## Acknowledgements

We are grateful to physicians and nurses at the acute respiratory infection clinic at Ramathibodi Hospital for their help in collecting the samples.

## Contribution of authors

EP, SPW, and AP designed the study, and wrote the manuscript. PJ, GW, and WS performed the study. KB and AP analysed the data. KB and SS edited the manuscript.

## Notes

### Competing Interest Statement

The authors have declared no competing interest.

### Clinical Trial

The study was not registered to any trial registry, as the study was not an interventional study.

